# Immunological Cross-Reactivity to Dengue Virus among Persons with Neuroinvasive West Nile Virus Infection

**DOI:** 10.1101/2022.01.03.22268686

**Authors:** Vanessa Raabe, Muktha S. Natrajan, Chris Huerta, Yongxian Xu, Lilin Lai, Mark J. Mulligan

## Abstract

Antibody dependent enhancement has been well described between Zika and dengue viruses, but is poorly characterized between West Nile and dengue viruses. We demonstrate that neuroinvasive West Nile virus infection leads to the development of non-neutralizing, cross-reactive IgG antibodies to dengue and Zika viruses capable of causing antibody dependent enhancement *in vitro* of dengue virus and leads to the formation of flavivirus cross-reactive memory B cells in some patients.

## Introduction

West Nile virus (WNV) is a single-stranded RNA virus in the Japanese encephalitis virus serocomplex of the *Flaviviriridae* family. Human infection is frequently asymptomatic although severe neuroinvasive disease, such as myelitis, meningitis, acute flaccid paralysis, and encephalitis, occurs in a minority of infected individuals [1]. After introduction to North America in 1999, WNV quickly spread throughout the continental United States leading to 21,574 reported neuroinvasive WNV infections between 1999 and 2016 [2].

Similar to other flaviviruses, antibodies generated to WNV primarily target the envelope (E) protein [3]. Cross-reactivity among certain flavivirus anti-envelope antibodies is well established and antibodies generated following dengue virus (DENV) immunization or Zika virus (ZIKV) infection demonstrate capacity to cross-react with WNV [4-6]. WNV antibodies are capable of binding to and mediating antibody-dependent enhancement (ADE) of ZIKV both *in vitro* and *in vivo*, a phenomenon by which non-neutralizing, cross-reactive IgG antibodies facilitate viral entry into Fc receptor bearing cells resulting in enhanced infection [7]. ADE of DENV following WNV infection has to our knowledge not been reported [7]. Here we report the development of non-neutralizing, cross-reactive IgG antibodies to DENV and ZIKV recombinant envelope proteins among three participants with neuroinvasive WNV infection and demonstrate potential for ADE of DENV.

## Methods

Patients with confirmed WNV neuroinvasive infection were identified and written informed consent for study participation was obtained from all patients or their next of kin under an Emory University Institutional Review Board-approved protocol for phlebotomy for diseases of public health importance.

Levels of WNV, DENV, and ZIKV IgG antibodies were assessed by Enzyme-Linked Immunosorbent Assay (ELISA) using described methods with WNV, DENV1-4, and ZIKV recombinant envelope (rE) proteins (MyBioSource #MBS5304480; CTK Diagnostics #A201 DENV1 VN/BID-V949/2007, #A2302 DENV-2 GWL39 IND-01, #A2303 DENV-3 US/BID-V/1090/1998 and #A2304 DENV4 321750; eEnzyme #ZV-E-005P) as target antigens [8]. Serum neutralization antibody assays were conducted via focus reduction neutralization tests (FRNT) to ZIKV (PRVABC59 strain, obtained from CDC), DENV1 (Hawaii strain, BEI resources NR-82), DENV2 (New Guinea C strain, BEI resources NR-84), DENV3 (Slemen 78 strain, donated by Jens Wrammert, Emory University), DENV4 (H241 strain, BEI resources NR-86), and Yellow Fever 17D (YF-VAX, Sanofi) as previously described with modification of the concentration of methylcellulose overlay to 1-2% wt/vol [9].

Antibody dependent enhancement was assessed with a protocol adapted from Bardina et al. by incubating K562 cells (ATCC, CCL 243) with 1 × 10^4^ focus-forming units of DENV2 for 48 hours in the presence of serial dilutions of heat-inactivated plasma from participants [7]. All assays were performed in duplicate. Plasma from a dengue-experienced person (SeraCare, 0325-0014) and a known flavivirus-naïve healthy control were used as positive and negative controls respectively. Briefly, cells were fixed and permeabilized (BD, 554722), incubated for 10 minutes with 1:500 Fc blocking antibody (Fisher Scientific, BDB564219), followed by staining with 1:1000 4G2 antibody (Millipore, MAB10216) for 1 hour. After washing, cells were incubated with goat anti-mouse 1:150 PE-IgG antibody (Columbia Biosciences, D5-112-100) for 1 hour in the dark, washed, and resuspended in 1% bovine serum albumin in PBS. Data was collected on a Cytoflex flow cytometer (Beckman Coulter). The percentage of DENV2 infected cells was averaged between duplicate runs; one exception to this was made for participant A at 1 × 10^3^ serum dilution in which one of the values was significantly elevated in a manner inconsistent with the duplicate result and other assays values and was subsequently excluded in final calculations. The average percentage of K562 cells infected for each participant was compared to the average percentage of infected cells across all negative control dilutions to calculate the fold increase of infection attributable to antibody dependent enhancement.

Memory B cell ELISpots were performed as previously described using WNV rE protein, DENV1-4 rE proteins, Yellow fever 17D virus, and ZIKV lysate [9].

## Results

Blood samples were collected from three patients with neuroinvasive WNV infection during 2017. Participants A and B presented with WNV encephalitis only while participant C developed both WNV encephalitis and myelitis. Participants A and B received intravenous immunoglobulin (IVIG) treatment during the acute phase of illness; participant C received corticosteroids and 5 days of plasmapheresis but did not receive IVIG. None of the participants reported a history of travel to DENV or ZIKV endemic regions or previous receipt of yellow fever virus (YFV) vaccination. Whole blood, serum, plasma, and peripheral blood mononuclear samples were collected on day post-symptom onset (DPO) 31 from participant A, DPO 32 and 146 from participant B, and DPO 138 from participant C. Participant B had near complete neurological recovery while participants A and C suffered from significant persistent weakness during convalescence.

All sera samples contained detectable IgG antibodies to WNV, DENV1-4, and ZIKV rE protein by ELISA. Calculated WNV endpoint titers were higher than DENV1-3 and ZIKV titers at all time points except DOI 32 for participant B while titers to DENV4 were higher than to WNV at all time points except for DOI 146 for participant B. WNV endpoint titers ranged from 522 to 8632, titers to DENV1-4 ranged from 265-11354, and ZIKV titers ranged from 335 to 2706 (Table 1). Low levels of neutralizing antibodies against DENV4 (FRNT_50_ 1:40) were detectable in participants A and C. Neutralizing antibodies were not detected (FRNT_50_ <1:20) against DENV1-3, ZIKV, or YFV-17D in participants A and C or to DENV1-4, ZIKV, or YFV in participant B at either time point (data not shown).

**Table 1.** Calculated ELISA Endpoint Titers to WNV and DENV serotypes in Three WNV-Infected Patients

|  | Participant A<br>DPO 31 | Participant B<br>DPO 32 | Participant B<br>DPO 146 | Participant C<br>DPO 138 |
| --- | --- | --- | --- | --- |
| WNV rE | 5900 | 522 | 7632 | 5632 |
| DENV1 rE | 265 | 918 | 580 | 337 |
| DENV2 rE | 2351 | 731 | 1132 | 805 |
| DENV3 rE | 636 | 2919 | 1825 | 339 |
| DENV4 rE | 11354 | 3762 | 2897 | 9763 |
| ZIKV rE | 2706 | 1940 | 335 | 1949 |

Antibody dependent enhancement of DENV2 infection was detectable at 1:10 plasma dilution for all participants (fold increase of infection: 3.6 for participant A, 3.4 for participant B at DPO 146, and 4.2 for participant C) and at 1:100 dilution for participant A only (6.6-fold increase of infection) [Figure 1]. No ADE of DENV2 infection was observed at the remaining plasma dilutions or at the DPO 32 time point for participant B.

**Figure 1.**
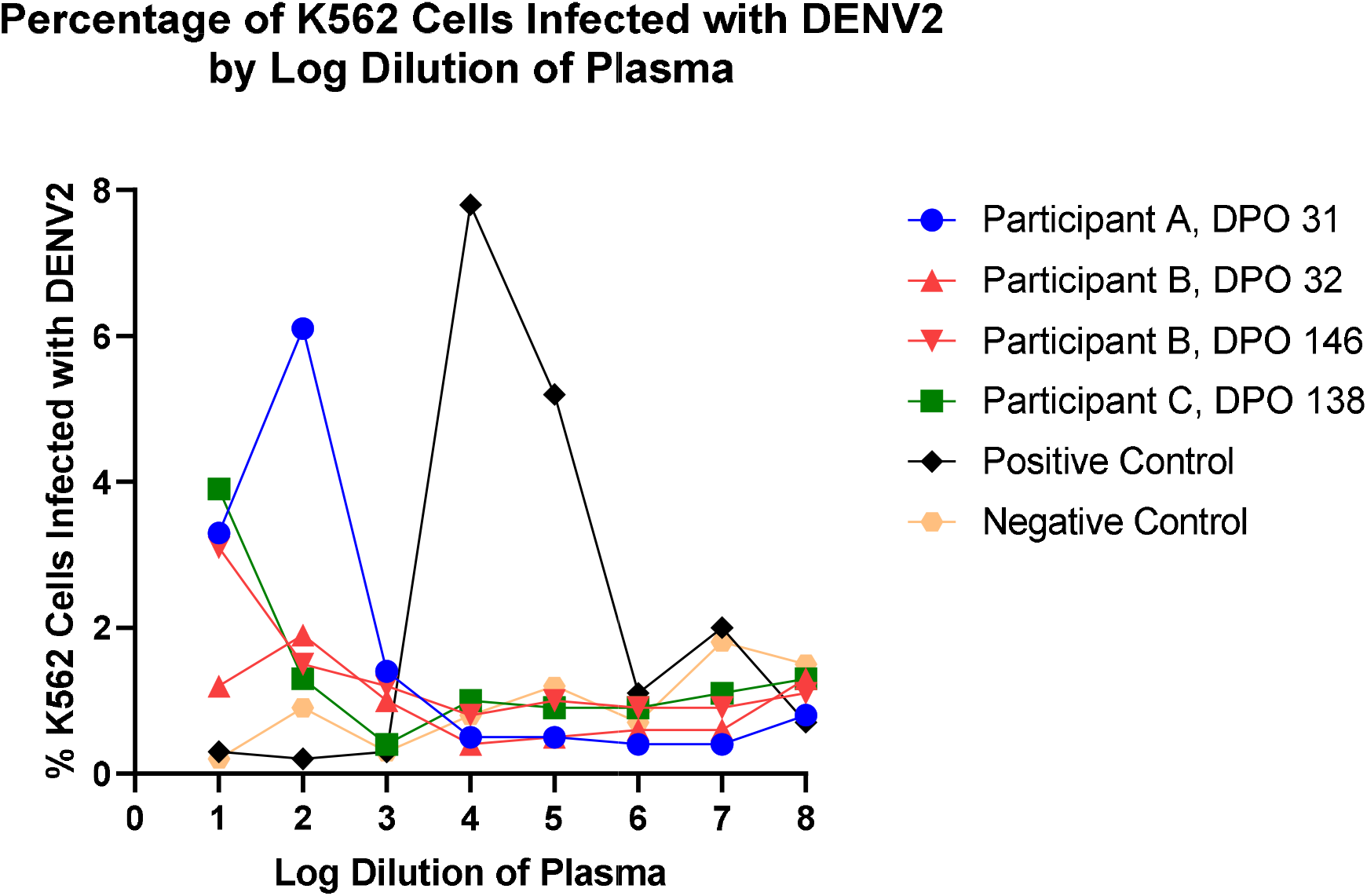
Percentage of K562 Cells Infected with DENV2 by Log Dilution of Plasma

Memory B cell (MBC) recognition of WNV rE was present in participant A (0.1% of IgG secreting MBCs) and participant C (0.36% of IgG secreting MBCs) but was not detectable at either time point for participant B. Flavivirus cross-reactive MBCs were observed in participant C to DENV3 (0.56% of IgG secreting MBCs) and ZIKV (0.04% of IgG secreting MBCs) [Supplementary Figure 1]. No flavivirus cross-reactive IgG-secreting MBCs were detectable in participants A and B.

## Discussion

Although WNV and DENV occupy separate antigenic complexes of the *Flaviviridae* family, cross-reactivity of WNV IgM and IgG antibodies with commercial assays for DENV antibodies has previously been demonstrated [5, 6]. Here we replicate the finding of cross-reactive IgG antibodies to recombinant envelope of all four DENV strains and ZIKV among three individuals with neuroinvasive WNV infection. Although two study participants received IVIG during the course of illness, the presence of cross-reactive IgG antibodies was detectable in the participant who did not receive IVIG treatment and persisted to DPO 146 in one of the IVIG treat participants, participant B. As DPO 146 occurred more than five times past the half-life of IVIG (25.8 days), the finding of cross-reactive IgG antibodies to DENV strains at this time point was unlikely to be solely secondary to IVIG administration [10]. One of the three participants demonstrated cross-reactive IgG-producing memory B cell responses to DENV3 and ZIKV, consistent with WNV infection eliciting an immune response capable of generating cross-reactive antibodies to other flaviviruses.

The potential for ADE of WNV following DNA immunization with DENV envelope protein domains I and II has been demonstrated in mice but not vice versa [4]. We demonstrated that the antibody response in three participants with neuroinvasive WNV infection has the capacity to cause ADE of DENV2 in K562 cells and likely other DENV serotypes not tested given antibody cross-reactivity by ELISA was observed to all four DENV serotypes. The WNV envelope protein is a potential antigen of interest in the development of WNV vaccines, therefore, antibodies generated following WNV vaccination could potentially cause antibody-dependent enhancement of DENV virus [11, 12]. We recommend that assessments for the development of cross-reactive antibodies to other flaviviruses, including DENV, should be investigated as a safety measure during WNV vaccine development.

## Data Availability

All data produced in the present work are contained in the manuscript.

## Acknowledgements

The authors would like to acknowledge the contributions of Joanne Altieri-Rivera, Alison Beck, Mary Bower, Tigisty Girmay, Vinit Karmali, Pamela Lankford-Turner, Dongli Wang, and Yerun Zhu from Emory University and Kristy Murray from Baylor College of Medicine.

## Footnotes

This research was conducted at the Hope Clinic of the Emory Vaccine Center at Emory University. Partial results included in the paper have previously presented in poster format at the NFID Clinical Vaccinology Course in November, 2018.

## Table and Figure Legends

**Supplementary Figure 1** Memory B Cell IgG ELISpot for Participants A-C to Various Flaviviruses

