## Supplementary figures and images for "Immunological Cross-Reactivity to Dengue Virus among Persons with Neuroinvasive West Nile Virus Infection"

### Supplemental Figure 1

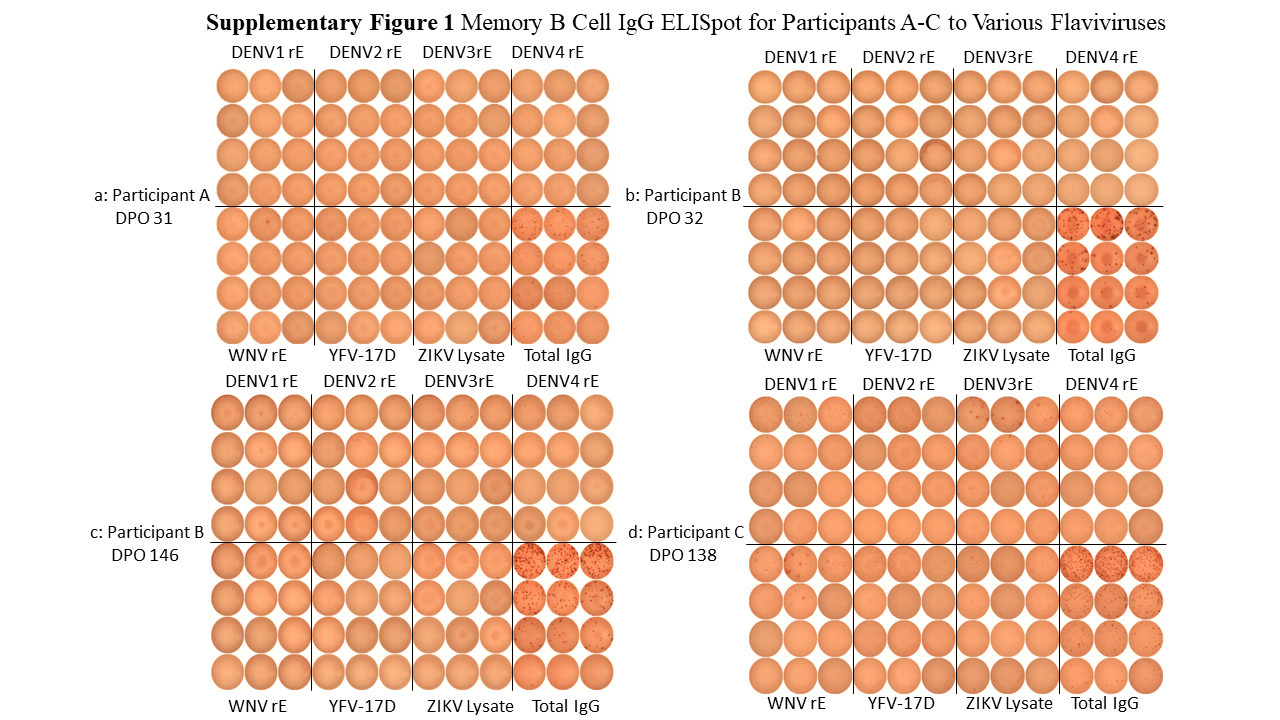
